# Discriminative Accuracy of CHA_2_DS_2_VASc Score, and Development of Predictive Accuracy Model Using Machine Learning for Ischemic Stroke in Cardiac Amyloidosis

**DOI:** 10.1101/2023.06.16.23291530

**Authors:** Waqas Ullah, Abhinav Nair, Eric Warner, Salman Zahid, Daniel R. Frisch, Indranee Rajapreyar, Rene J. Alvarez, Mohamad Alkhouli, Sridhara S Yaddanapudi, Mathew S. Maurer, Yegeny Brailovsky

## Abstract

**Background:** Cardiac amyloidosis (CA) in conjunction with atrial fibrillation (AF) presents unique management challenges. CHA2DS2VASc score in these patients is believed to underestimate the risk of ischemic stroke, necessitating a better predictive model in these patients.

**Methods:** Data was obtained from the National Readmission Database (NRD). Outcomes between CA-AF and no-CA-AF were compared using multivariate regression analysis to calculate adjusted odds ratios (aOR). AutoScore; an interpretable machine learning framework, was used to develop a stroke risk prediction model, the predictive accuracy of which was evaluated with an area under the curve (AUC) using the receiver operating characteristic analysis.

**Results:** A total of 11,860,804 (CA-AF 22,687 [0.19%] and no-CA-AF 11,838,117) patients were identified from 2015-2019. The adjusted odds of mortality (aOR 1.41 and 1.29), stroke (aOR 1.78 and 1.74), non-intracranial hemorrhage (aOR 2.10 and aOR 1.85), and intracranial hemorrhage (aOR 14.4 and aOR 4.26) were significantly higher in CA-AF compared with non-CA-AF at both index admission and 30-days, respectively. The CHA_2_DS_2_VASc score had a poor discriminative accuracy for stroke at 30-days in CA-AF (AUC 49%, 95%CI 47%-51%, p=0.54). The machine learning autoscore integrative model revealed that the predictive ability of our newly proposed E-CHADS score (end-stage renal disease (ESRD), congestive heart failure, hypertension, active cancer, dementia, and diabetes mellitus) for 30-day risk of ischemic stroke in CA-AF was excellent (for a cutoff of 52 points random forest score) with an AUC of 80% (95%CI 74%-86%)

**Conclusion:** Cardiac amyloidosis carries a high risk of ischemic stroke that is not accurately predicted by the CHA_2_DS_2_VASc score. Our proposed model (E-CHADS) identifies 3 new variables (ESRD, dementia, and cancer) that have higher discriminative accuracy for ischemic stroke in these patients.

## INTRODUCTION

Atrial fibrillation (AF) significantly burdens society and the healthcare system. The current prevalence of AF in the United States (US) is about 5.2 million, estimated to rise to 12.1 million by 2030 [1]. Despite strong efforts by professionals, and the scientific community, AF remains a leading cause of heart failure (HF) related hospitalizations, ischemic stroke, and mortality [1]. It is the most commonly encountered arrhythmia in patients with HF, particularly those with cardiac amyloidosis (CA) (in ∼70% of patients) [2]. The association of AF and CA has important clinical implications. AF-related loss of atrial contribution to an already hypertrophied and dysfunctional left ventricle is often poorly tolerated, which not only results in profound clinical deterioration but also leads to recurrent hospitalizations [3]. CA-associated atrial dilatation exponentially increases the risk of intracardiac thrombus, systemic embolism, and ischemic strokes [4,5].

Given the complex interplay of AF and CA, the traditional risk assessment tool (CHA_2_DS_2_VASc score) poorly estimates the risk of ischemic stroke [6]. Some data recommends the use of oral anticoagulation (OAC) in CA-AF patients regardless of the CHA_2_DS_2_VASc score and even after restoration of the sinus rhythm [6, 7]. However, there has been no study to assess the accuracy of CHA_2_DS_2_VASc, and to determine a better predictive risk assessment tool for ischemic stroke in patients with CA-AF. Providers are often left to expert consensus and clinical experience to aid in decision-making. The current study aimed to assess the thrombotic and bleeding outcomes of CA-AF in the context of OAC and to evaluate the discriminatory accuracy of the CHA_2_DS_2_VASc score. Using a machine learning algorithm, this study also sought to develop a model that had a better predictive ability for ischemic stroke.

## METHODS

### Data Source

Data was obtained from the Nationwide Readmissions Database (NRD) which is part of the Healthcare Cost and Utilization Project. It was established by the federal-state-industry partnership and monitored by the Agency for Healthcare Research and Quality. The NRD is an all-payer database that contains data from more than 18 million discharges from 22 US states each year. The unweighted data of NRD accounts for 49% and 51% of the total US hospitalizations and population, respectively. NRD contains unique identification codes that can link patients across the same year, allowing us to capture readmission. Data is anonymized and hence exempted from the approval of the institutional review board (IRB).

### Study design and population

Using the International Classification of Diseases-10th Revision-Clinical Modification [ICD-10-CM] codes, all hospitalizations for AF were identified between September 2015 and November 2019. (Supplementary [S.] Table S1) As data is annualized, hospitalizations in December from each year were excluded, to enable 30-day outcomes for each index admission. Using the standard ICD-10 code for organ-specific amyloidosis and cardiac failure, patients with cardiac amyloidosis were identified. All patients with AF were divided into two groups, those with CA and without CA. Each group was stratified based on the risk of ischemic stroke determined by the CHA_2_DS_2_VASc (congestive heart failure, arterial hypertension, age ≥75 years, diabetes mellitus, stroke, vascular disease, age 65–74 years, sex). The CHA_2_DS_2_VASc score was categorized into three groups; low risk (CHA_2_DS_2_VASc=0-1), moderate risk (CHA_2_DS_2_VASc=2), and high risk (CHA_2_DS_2_VASc≥3).

### Study Objectives and Outcomes

The major objectives and outcomes of our study can be categorized into three major sections. 1. Comparing the index-admission and 30-day adjusted risk of in-hospital mortality, ischemic stroke, non-intracranial hemorrhage (non-ICH), and ICH between patients with CA vs. without CA; stratified by CHA_2_DS_2_VASc score and long term use of OAC. 2. Assessing the discrimination accuracy of CHA_2_DS_2_VASc score for 30-day ischemic stroke in patients with AF and concomitant diagnosis of CA. 3. Developing a predictive model with a machine learning algorithm for the risk of ischemic stroke in patients with AF and CA.

### Statistical analysis

#### Outcomes of CA-AF vs. non-CA-AF

Categorical variables were summarized as percentages and frequency and compared using the chi-squared test. The proportion of random missing data was <1% and hence was excluded. A binomial multivariable logistic regression model was used to estimate adjusted odds ratios (aOR) for mortality, stroke, ICH, and non-ICH. Net results were further stratified based on the CHA_2_DS_2_VASc score and history of prior use of OAC therapy. A p-value interaction analysis enabled an evaluation of the impact of potential effect modifiers. The cumulative incidence of major outcomes was assessed using the Kaplan Meier (KM) curves.

### Discrimination Accuracy of CHA_2_DS_2_VASc Score for Ischemic Stroke in CA-AF

Next, the performance of the CHA_2_DS_2_VASc risk score and CHA_2_DS_2_VASc categories in predicting the risk of ischemic stroke across CA-AF was determined. An area under the curve (AUC) was calculated using receiver operating characteristic (ROC) analysis, to assess how well the logistic regression model fits the dataset.

### Development of Machine Learning Predictive Model for Ischemic Stroke in CA-AF

Finally, a stroke prediction model was created using the set guidelines by TRIPOD (Transparent Reporting of a multivariable prediction model for Individual Prognosis Or Diagnosis). [8] The final 30-day readmission data was divided into three non-overlapping cohorts in a proportion of 70%, 20%, and 10% for training-, validation- and testing-sets, respectively. The continuous variables in the dataset were transformed into categorical variables with the cut-off values determined by the quantiles of the data points.

A combined regression and machine learning approach was used to create a predictive accuracy model. For the latter, an AutoScore framework was utilized to automate the derivation of point-based risk scores (range 1-100) for variables that had a biologically plausible relation with the outcome (ischemic stroke) [9, 10]. A total of 42 variables based on clinical relevance comprising components of the CHA_2_DS_2_VASc score, demographics, and major baseline comorbidities were fed into the model as potential predictors. (Table S14) Random forest (RF), a well-validated machine learning algorithm, was utilized to rank the variables in order of their importance for ischemic stroke prediction [11]. The RF model is a non-parametric method and consists of multiple tree-structured classifiers which evolve to a maximum size by training on a random selection of variables and a bootstrap sample. The RF ranking is based on the virtue of variables being heterogeneous and the absence of nonlinearity, which provides an advantage over LASSO regression methods to allow for the optimal selection of variables for risk prediction.

A multivariable logistic regression model was used to obtain a coefficient value for score weighting. Among the variables, the lowest coefficient was selected as our reference to eliminate variables with a negative coefficient value. A parsimony and AUC plots were generated illustrating cumulative RF-ranking scores of variables and model performance, respectively. A variable at the plot’s tail-end signified that adding the variable to the model will not improve the predictive accuracy.

For the optimal predictive model, multiple iterations and fine-tuning enabled a selection of the best cut-off value of the RF-ranking score for each variable, and choose the final number of variables in the model. To enhance the interpretability of the model, the RF-ranking score was converted into a probability (as shown by AUC), using weighted logistic regression analysis on the validation set. The prediction performance (accuracy, sensitivity, specificity, negative and positive predictive value (NPV, PPV) of the model was computed using the cumulative score cut-off values of the included variables against the risk of ischemic stroke. The predictability and performance of the model were assessed using the ROC analysis, and AUC with its 95% confidence interval. A two-sided p < 0.05 was considered statistically significant. Lastly, the testing set that was blinded to the scoring algorithm evaluated the performance of the model. Data characteristics and adjusted estimates of (no ischemic stroke vs. ischemic stroke) based on each predictor in the model were also reported. The model was built using the package “AutoScore”, and “pROC”, all analyses were performed on R version 4.0.2.

## RESULTS

### Selection of Cases

From 2015 to 2019, a total of 11,860,804 weighted samples of AF patients were identified at index admission. Of these, 11,838,117 (99.8%) had no history of CA, while 22,687 (0.2%) had a diagnosis of CA. The flow diagram of the selection of cases is presented in Figure S1. The ICD-10 codes used to identify cases and characteristics are presented in Table S1.

### Baseline Characteristics

The detailed intergroup comparison of demographics and key comorbidities is presented in Table S2. In both non-CA (87.1%), and CA (92.1%), the predominant CHA_2_DS_2_VASc designation was high risk, mostly having CHA_2_DS_2_VASc class 4 (27.2% vs. 32.7%, respectively). (Table S3, Figure S2) The most common contributors to the CHA_2_DS_2_VASc score were a history of hypertension (87.8% vs. 84.1%), followed by age >75 years (65.5% vs. 68.3%). (Figure S3) Only 35.9% of the total population with CA-AF were on long-term OAC therapy at the index admission. As the CHA_2_DS_2_VASc risk category increased from low to moderate to high, the proportion of anticoagulation use increased from 18.0% to 31.7% to 35.9%, respectively. (Figure S4) The latter percentage increased to 37.1% at 30-day readmission. (Table S4) On a yearly trend analysis, the annual index admission rate for patients with CA-AF per total CA-AF in the study increased significantly from 18.4% in 2016 to 30.6% in 2019. (Table S5, Figure S5)

### Overall Outcomes at Index Admission and 30-Days

Patients with CA-AF (vs. non-CA-AF) had significantly higher unadjusted odds of all major complications. Similarly, the adjusted odds of mortality (aOR 1.41, 95% CI 1.34-1.48 and aOR 1.29, 95% CI 1.19-1.40), stroke (aOR 1.78, 95% CI 1.68-1.89 and aOR 1.74, 95% CI 1.56-1.94), non-ICH major bleeding (aOR 2.10, 95% CI 1.97-2.24 and aOR 1.85, 95% CI 1.67-2.17), and ICH (aOR 14.4, 95% CI 13.91-15.01 and aOR 4.26, 95% CI 3.83-4.73) remained significantly higher in CA-AF compared with non-CA-AF at both index admission and 30-days, respectively (Tables S6, Figure 1).

**Figure 1:**
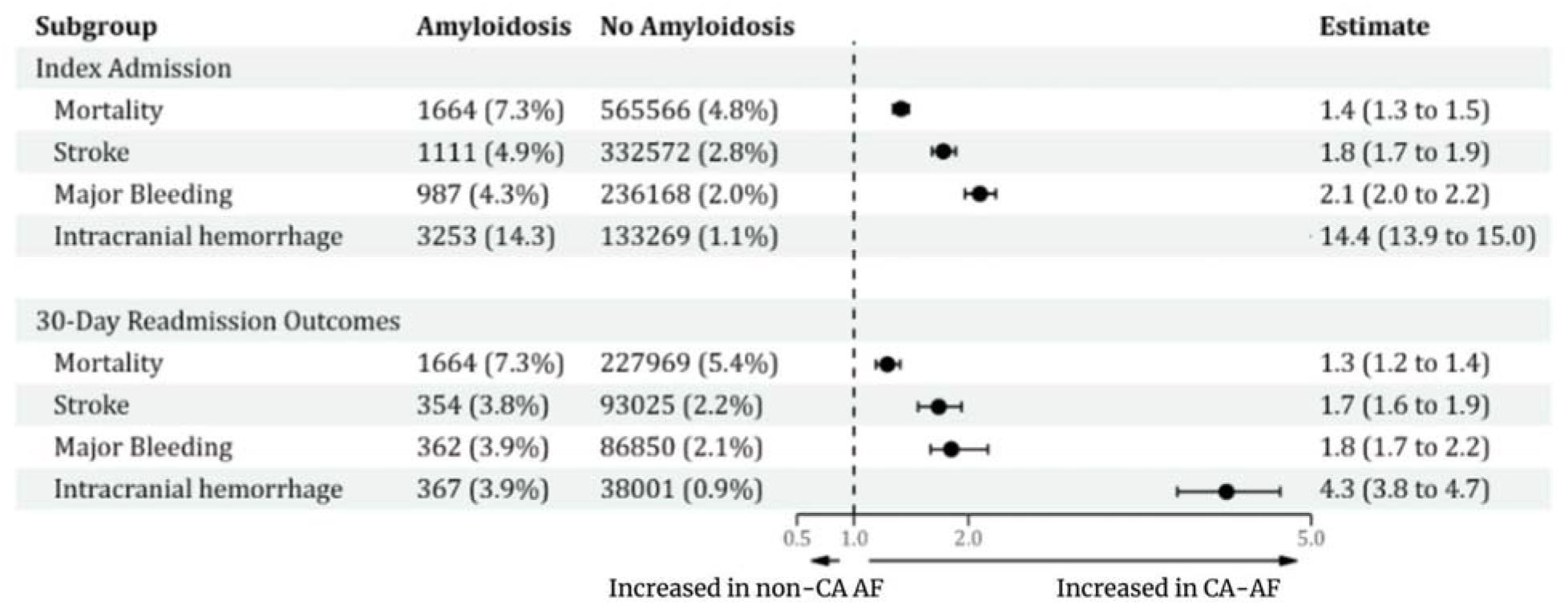
Proportion of major events and estimates of outcomes between CA-AF and no-CA-AF at index admission and 30-day readmission

### Adjusted Index Admission and 30-Day Outcomes stratified by the CHA_2_DS_2_VASc Risk and Long-term Anticoagulation Use

The stratified frequency and estimates of outcomes are given in Tables S7-S8. In concordance with the overall outcomes, the CA-AF group remained to have a higher risk of mortality, ischemic stroke, non-ICH, and ICH across all CHA_2_DS_2_VASc categories and irrespective of the use of OAC at both index-admission and 30-days. Except that, anticoagulation use in medium (aOR 0.64 95% CI 0.29-1.41 and aOR 0.74, 95% CI 0.27-1.99) and high risk (aOR 1.56, 95% CI 0.74-1.90 and aOR 1.18, 95% 0.94-1.47) patients with CA-AF vs. non-CA-AF had a non-significantly different incidence of ischemic stroke at index admission and 30-days, respectively. OAC did not significantly increase the risk of ICH in CA-AF. Moreover, CA was independently associated with a higher risk of stroke even in the absence of AF. (Table S9) Interaction analysis showed that patients not on OAC had a significantly higher risk of stroke in CA-AF compared with both CA-AF on OAC and those without CA-AF, at index admission as well as 30-days. (Table S10, Figures S6-S7)

### Receiver Operating Characteristics of the CHA_2_DS_2_VASc Score and Risk Category for Ischemic Stroke

In CA-AF patients, both CHA_2_DS_2_VASc score (AUC 0.493, 95% CI 0.470-0.516, p=0.54) and CHA_2_DS_2_VASc risk category (AUC= 0.509, 95% CI 0.487-0.532, p=0.423) had poor discriminatory accuracy for ischemic stroke (Table S11, Figure S8).

### Machine Learning and Predictive Model Development Score development

45 variables were used to generate a ranking list in the predictive model. After adjusting for coefficient and fine-tuning the model, a scoring table ranging from 0-100 was obtained. Using the autoscore algorithm, the RF-scores were ranked in order of their importance (Figures S9, S10). A parsimony plot was generated based on the predictive ability of the RF-score (Figure 2) The top 7 variables that achieved the maximum AUC were selected in the final model.

**Figure 2.**
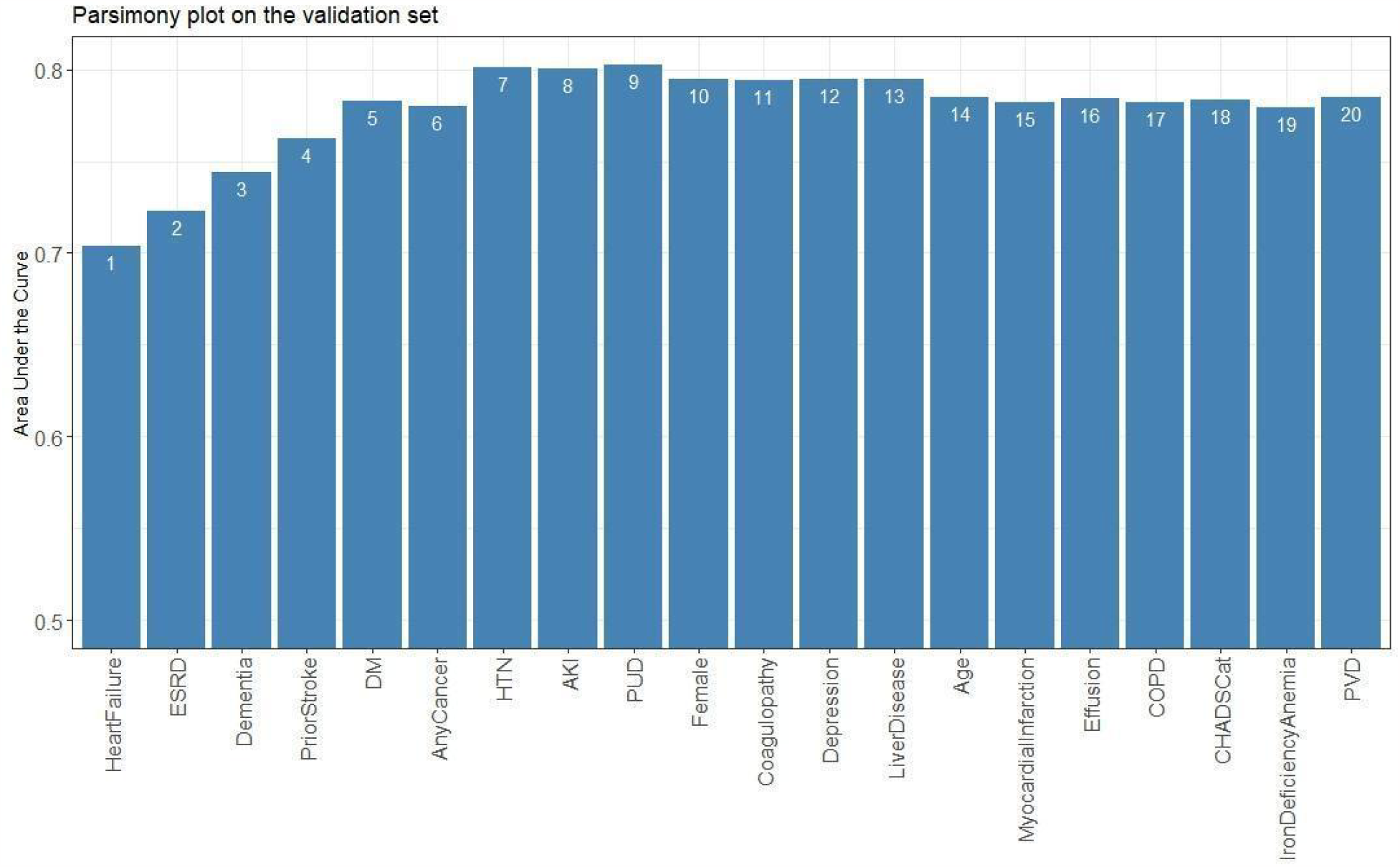
Parsimony plot of the new model’s performance predicting stroke (as indicated by the area under the curve) as a function of the model’s complexity (as noted in the number of variables included in the model). The AUC (accuracy of the model) increased with the addition of variables till HTN; no further increase in accuracy could be achieved with the addition of other variables beyond variable 8 (AKI). This indicates that the model’s highest accuracy was attained with the first seven variables.

A higher score indicated a higher risk of ischemic stroke. HF (23), HTN (19), ESRD (19), and prior stroke (15) were associated with the highest likelihood of ischemic stroke, followed by dementia (8), DM (4), and cancer (4). (Table S12) Addition of other variables or components of the CHA_2_DS_2_VASc score (age, female sex, vascular disease) did not increase the predictive ability of the model. (Figure 2) For a specific score cut-off threshold, the probability of a 30-day ischemic stroke (predicted risk), and the accompanying metrics (accuracy, sensitivity, specificity, NPV, and PPV) were recorded in Table 1. For specific probability thresholds, their respective score cutoffs were recorded in Table S13. At a threshold cut-off >52, our proposed E-CHADS model performance was the best with sensitivity of 0.71 (95% CI: 0.54-0.88), specificity of 0.77 (95% CI: 0.73-0.81) and AUC of 0.80 (95% CI: 0.74-0.86). (Figure 3, S11) Finally, the adjusted odds of having an outcome for each of the potential predictors that were fed in the model are shown in Table S14, which confirms a higher risk of stroke with the 7 selected components of the E-CHADS.

**Table 1.** Model performance at specified score cutoffs and predicted risk of stroke

| Score cut-off | Predicted Risk | % of patients | Accuracy (95% CI) | Sensitivity (95% CI) | Specificity (95% CI) | PPV (95% CI) | NPV (95% CI) |
| --- | --- | --- | --- | --- | --- | --- | --- |
| ≥20 | ≥0.80% | 95 | 9.8 (7.9-11.7) | 100 (100-100) | 5.5 (3.6-7.5) | 4.8 (4.7-4.9) | 100 (100-100) |
| ≥40 | ≥2.40% | 55 | 48.9 (44.7-53.2) | 95.8 (87.5-100) | 46.6 (42.3-51.2) | 7.9 (7-8.7) | 99.6 (98.7-100) |
| ≥60 | ≥5.90% | 22 | 79.2 (75.8-82.6) | 66.7 (49.9-83.3) | 79.8 (76.3-83.2) | 13.6 (9.7-17.5) | 98.1 (97-99) |
| ≥75 | ≥11.60% | 9 | 88.5 (86-90.8) | 20.8 (4.2-37.5) | 91.7 (89.3-94.1) | 10.4 (2.9-18.9) | 96 (95.3-96.9) |
| ≥90 | ≥21.60% | 2 | 94.3 (93.2-95.3) | 4.2 (0-12.5) | 98.6 (97.6-99.6) | 11.1 (0-42.9) | 95.6 (95.4-96) |
| ≥100 | ≥29% | 0 | 95.5 (94.9-96) | 4.2 (0-12.5) | 99.8 (99.4-100) | 50 (0-100) | 95.6 (95.4-96) |

**Caption:** Machine learning model's predicted risk of stroke based on scores determined by 8 selected variables (heart failure, ESRD, HTN, Neurological disease, dementia, electrolyte abnormalities, DM, and any cancer). Also shown is the accuracy, sensitivity, specificity, PPV, NPV, and their respective 95% CI's for the model at each score cut-off point.
**Abbreviations:** ESRD: end-stage renal disease, HTN: hypertension; DM: diabetes mellitus, PPV: positive predictive value; NPV: negative predictive value, CI: confidence interval
**Note:** Values for accuracy, sensitivity, specificity, PPV, NPV, and their respective confidence intervals are listed as percentages

**Figure 3.**
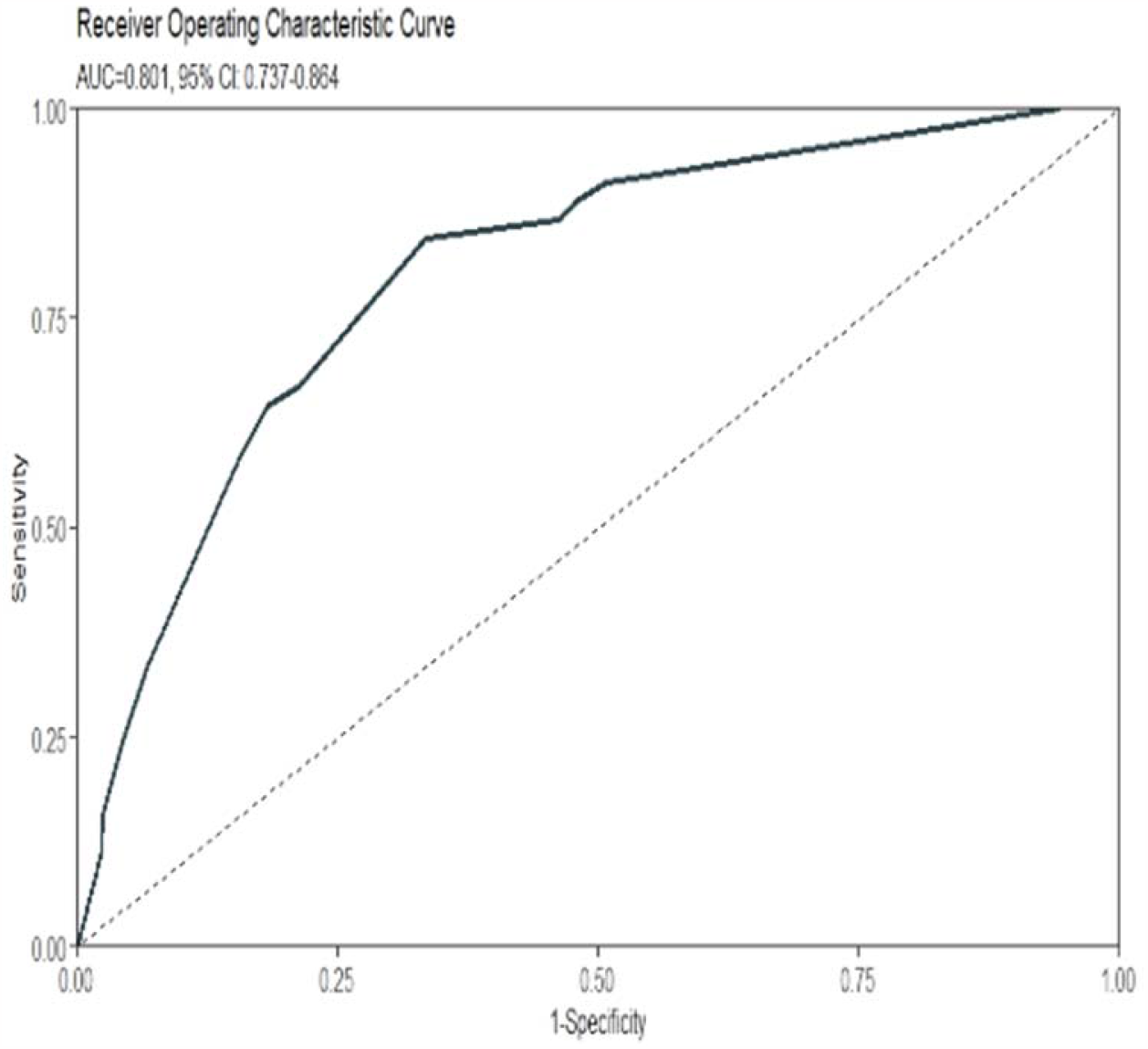
ROC Curve for E-CHADS Model showing a high AUC at 80% for a cut-off RF score of 52 for identifying 30-day risk of ischemic stroke in patients with CA-AF.

### Performance evaluation

The testing data set was utilized to assess the E-CHADS prediction performance. The proposed model achieved similar sensitivity and specificity, with a high discrimination power of AUC 0.79 (95% CI: 0.71-0.86).

## DISCUSSION

The current study is the first to compare outcomes, assess the discriminative power of the CHA_2_DS_2_VASc score, and develop the optimal predictive model (E-CHADS) for ischemic stroke in patients with CA and concomitant AF. The major findings are summarized as follows; 1) patients with CA-AF had a significantly increased adjusted risk of mortality, ischemic stroke, non-ICH, and ICH at index admission of AF and 30-day follow-up. 2) The higher risk of non-ICH and ICH in CA-AF was independent of the use of OAC and CHA_2_DS_2_VASc risk category, however, the incidence of ischemic stroke attenuated by 1.57 times with the use of anticoagulation, especially in medium and high-risk patients with CA-AF. 3) CHA_2_DS_2_VASc score had a poor discriminative accuracy for ischemic stroke at 30 days reaching up to only 50% AUC. 4) Our proposed E-CHADS model, by adding ESRD, active cancer, and dementia with the four components of CHA_2_DS_2_VASc score (HF, HTN, DM, prior stroke) exhibited excellent discrimination capability by increasing the predictive accuracy of a stroke to 80% (at a threshold of RF-score >52).

Our study showed that CA was independently associated with a higher risk of in-hospital ischemic stroke [6]. Patients with CA had a 5% risk of stroke even in absence of AF, compared with 2.5% in patients with AF without CA. This could be attributed to a heightened risk of cardiac thrombus formation, endothelial injury, coagulation pathway defect, hyposplenism-induced thrombocytosis, and nephrotic syndrome that are frequently associated with CA [12]. Despite an ∼50% (statistically significant) reduction in the risk of ischemic stroke with OAC, only 37% of the total patients with CA-AF were on anticoagulation at 30-days of index AF, 92.8% of whom belonged to the high-risk, 6.5% to moderate and only ∼1% to low-risk category. This not only indicates the efficacy of OAC in these patients but also highlights the fact that the decision to start OAC was probably based on the CHA_2_DS_2_VASc score, which we found to be a very poor predictor of stroke. These findings are supported by a recent Vilches et al. study that also found a substantially increased risk of systemic embolism in non-anticoagulated patients with AF and CA [7]. Furthermore, that study also demonstrated CHA_2_DS_2_VASc to be a poor predictor of systemic embolism [7]. Our study confirms these findings and thus identifies the unmet need for better awareness and advocacy for OAC therapy in all patients with CA-AF irrespective of the CHA_2_DS_2_VASc score to achieve the optimal antithrombotic benefits.

The biggest fear around OAC use in the clinical community is the augmented risk of major bleeding in CA. The current study showed a 21% higher risk of ICH in absence of AF, and up to a 12-fold increased odds of ICH compared with no-CA-AF (Figure S8, S9). However, it is important to note that this bleeding might be due to the inherent nature of CA-induced vascular fragility, clotting abnormalities, and concomitant cerebral amyloid angiopathy rather than the use of OAC therapy, as the risk of bleeding in CA-AF remained invariantly high even in patients, not on AC. In the p-value interaction analysis, the antithrombotic benefits of anticoagulation seemed to outweigh the potential risk of bleeding as indicated by a decrease in the risk of ischemic stroke, but no increase in ICH with OAC use. Nonetheless, together, these findings underscore the importance of careful evaluation, judicious use of anticoagulation, individualization of care, and assessment of the bi-risk nature of CA in patients with AF.

Due to the poor predictive ability of the CHA_2_DS_2_VASc score, we developed a predictive model (E-CHADS) for ischemic stroke using a machine learning algorithm coupled with a well-validated random forest regression analysis. Four of the E-CHADS variables (HF, HTN, DM, and prior stroke) were components of the CHA_2_DS_2_VASc score, while we identified 3 new variables: ESRD, dementia, and active cancer. The proposed components of E-CHADS carry a strong biological plausibility in the pathogenesis of stroke. Studies have shown that HF increases the risk of ischemic stroke through the cardioembolic phenomenon, accumulation of traditional risk factors, and higher prevalence of permanent AF [12]. Similarly, HTN and DM have both been shown to induce vasculopathy by endothelial dysfunction, vascular remodeling, and inflammation; leading to an increased risk of stroke [13, 14]. For their part, cancer and ESRD induce a hypercoagulable state through the release of thrombogenic substances such as extracellular vesicles and factor X [15]. The latter also accentuates the activity of clotting factors due to the loss of function of antithrombin factors among other mechanisms [16]. Patients with vascular dementia and prior stroke possibly provide a substrate for future ischemic cerebrovascular events [17]. The point-based scoring structure in our model not only identifies the role of these factors but also ranks their relative importance in the model, which corresponds with clinical intuition. For instance, HF and HTN had the highest weightage on the model compared with dementia indicating that the former two are the most important variables in estimating the risk of stroke.

Overall, the findings of the current study have important clinical implications. Our study affirms that the CHA_2_DS_2_VASc score in its totality is a poor predictor of stroke in AF patients with concurrent CA. Consistent with the emerging notion, the female sex was found to be not a strong predictor of stroke [18]. Some components of CHA_2_DS_2_VASc (CHF, HTN, DM, and prior stroke) had a strong predictive power when combined with our newly proposed three variables (ESRD, active cancer, and dementia). Given the paucity of evidence on a predictive score for stroke, and the higher prevalence of AF in CA, our proposed model of E-CHADS reinforces the identification of key comorbidities, where simple addition of the RF-score in the model can predict the risk of ischemic stroke. Since it is easily accessible and readily obtainable, physicians could choose a cut-off tailored to their applications based on the likelihood of a stroke and metrics such as sensitivity and specificity. Whether this score helps in the decision-making for anticoagulation therapies requires future randomized studies.

## LIMITATIONS

Given the retrospective observational study design, there is a possibility of selection bias and residual confounding. For the same reason, we could not establish a causal relationship, but could only report temporal associations between CA and outcomes. Despite regression-based adjusted analysis, the impact of unmeasured covariates could not be determined. NRD is a readmission database linked to inpatient discharge records, so we could not capture events occurring outside the hospital and could not account for the competing risk of readmission, such as mortality occurring in the community or ambulatory settings. Furthermore, the lack of data on disease severity, type of amyloidosis, long-term follow-up data, echocardiographic parameters, functional status, and medication use precluded our ability to perform a more robust stratified analysis, or include them in our machine learning model. Although all codes were verified using the standard recommended sources and with reported prior literature, the possibility of inadvertent coding error due to the lack of coding precision could not be entirely excluded. The diagnostic workup of CA is not widely available, so it is plausible that the perceived low incidence of CA, may be due to underdiagnosis or under-coding.

## CONCLUSION

CA is independently associated with a high risk of ischemic stroke and intracranial hemorrhage, the former being not predicted by the CHA_2_DS_2_VASc score. Anticoagulation lowers the risk of ischemic stroke without further increasing the incidence of major bleeding. We propose an E-CHADS score, a readily accessible risk prediction tool for ischemic stroke probability estimation in patients with CA-AF. Compared with the CHA_2_DS_2_VASc score, it presents the best performance that can help in the identification of high-risk patients. Future large-scale prospective studies are needed to validate our findings.

## PERSPECTIVES

### COMPETENCY IN MEDICAL KNOWLEDGE

Cardiac amyloidosis is independently associated with a higher risk of thrombosis and intracranial hemorrhage. The discriminative accuracy of the CHA_2_DS_2_VASc score for stroke is very poor in these patients.

### TRANSLATIONAL OUTLOOK

Further studies are needed to validate the predictive ability of our proposed E-CHADS model for ischemic stroke in patients with cardiac amyloidosis.

## Data Availability

Not publicly available

## Acknowledgments

None.

---

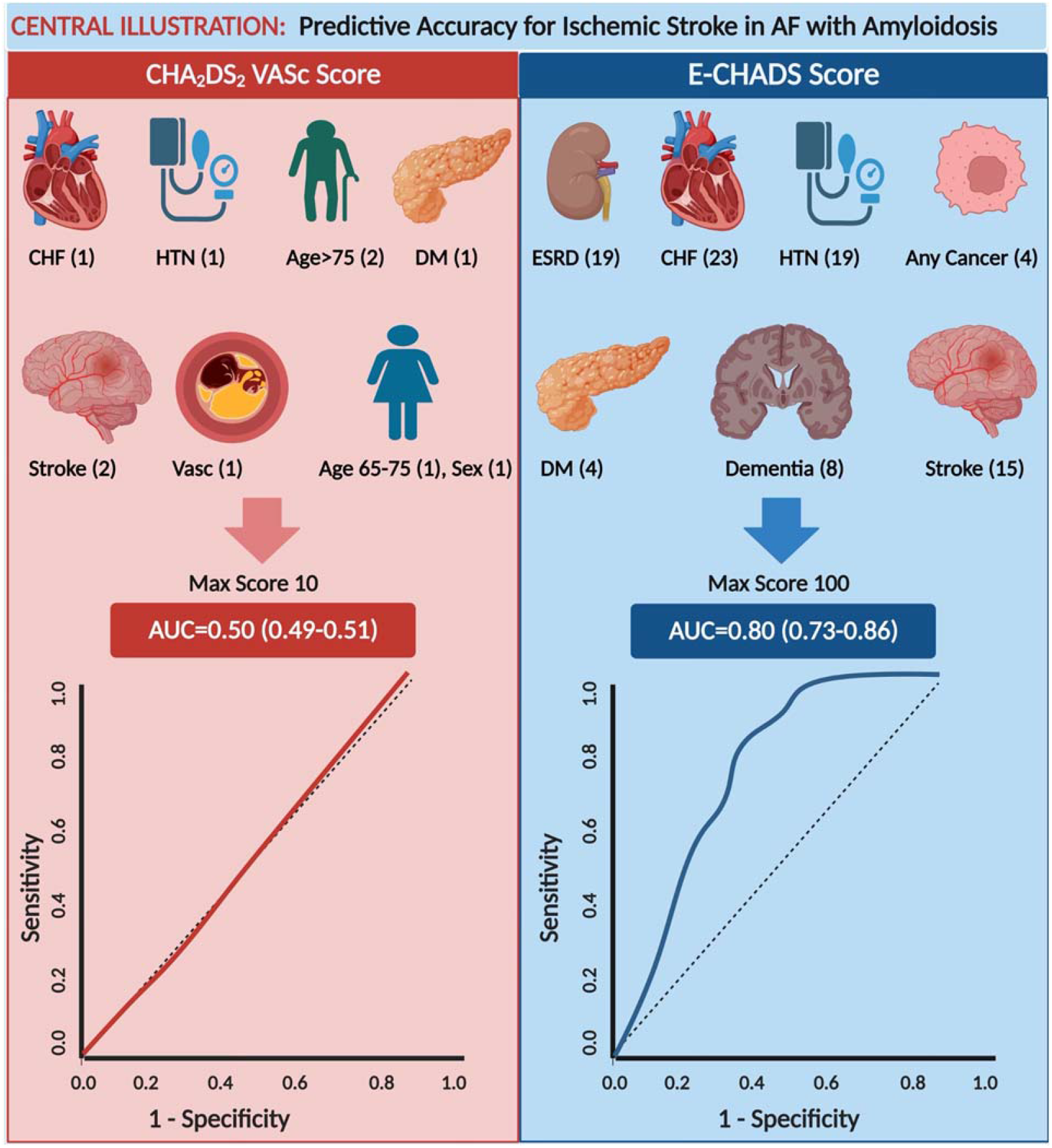
Central Illustration. The CHA_2_DS_2_VASc model (on the left) was a poor predictor of stroke in AF patients with comorbid cardiac amyloidosis, with an AUC = 0.50. The new model E-CHADS Score (on the right) appreciably improves in predictive accuracy for stroke in AF patients with cardiac amyloidosis, with an AUC = 0.80. Variables in each model’s scoring system are listed with their individual random forest scores; the high cumulative score represents higher risk of ischemic stroke in CA-AF at 30-day readmission. **Abbreviations:** CHF – congestive heart failure, HTN – hypertension, DM – diabetes mellitus, Vasc – vascular disease, AUC: area under the curve

## Notes

### Competing Interest Statement

The authors have declared no competing interest.

### Funding Statement

No external funding received

### Author Declarations

Patient data came from publicly available articles

